# How many relevant SARS-CoV-2 variants might we expect in the future?

**DOI:** 10.1101/2021.11.17.21266463

**Authors:** Roberto Littera, Maurizio Melis

## Abstract

**Objectives:** The emergence of new SARS-CoV-2 variants is a major challenge in the management of Covid-19 pandemic. A crucial issue is to quantify the number of variants which may represent a potential risk for public health in the future.

**Methods:** We fitted the data on the most relevant SARS-CoV-2 variants recorded by the World Health Organization (WHO). The function exploited for the fit is related to the total number of infected subjects in the world since the start of the epidemic.

**Results:** We found that the number of new relevant variants per ten million cases diminished by 30.4% between March 2020 and March 2022 (from 1.25 to 0.87). However, the decrease is now very slow and a further reduction by 10% would happen only for 5.6 billion infections in the world, i.e. ten times the cases from the beginning of the epidemic up to June 2022.

**Conclusion:** Our simple mathematical model can provide an estimate of the number of relevant SARS-CoV-2 variants as the cumulative number of cases increases worldwide and may represent a useful tool in planning strategies to effectively contrast the pandemic.

## Introduction

Most mutations in the genome of the severe acute respiratory syndrome coronavirus (SARS-CoV-2) are neutral or only mildly deleterious. However, a small proportion of mutations can increase infectivity and promote virus-host interactions that are critical to the establishment of persistent and more severe infection [1, 2]. For example, mutations in the spike protein, which mediates attachment of the virus to host cell-surface receptors [3], can have significant effects on virus behaviour. In order to effectively control the pandemic, it is imperative to investigate the emergence and spread of variants with an impact on disease transmission and human health [4].

SARS-CoV-2 sequences are shared daily on public databases such as the Global Initiative on Sharing All Influenza Data (GISAID) [5] or the European Centre for Disease Prevention and Control (ECDC) [6, 7], which significantly contribute to surveillance of the pandemic.

The World Health Organization has established that SARS-CoV-2 variants representing a possible risk to public health can be divided into three distinct categories [1]: variants under monitoring (VUMs), variants of interest (VOIs) and variants of concern (VOCs).

*Variants Under Monitoring* (VUMs) are associated with genetic mutations which alter virus characteristics, although evidence of phenotypic or epidemiological impact is still unclear.

*Variants of Interest* (VOIs) are associated with: *i)* genetic mutations which affect transmissibility, disease course, diagnostic or therapeutic escape; *ii)* relevant community transmission with an emerging risk to global public health.

*Variants of Concern* (VOCs) are associated with one or more of the following characteristics: *i)* increase in transmissibility; *ii)* increase in virulence or change in disease severity; *iii)* decrease in effectiveness of social measures, diagnostics, vaccines and therapeutics.

Given the continuous evolution of coronavirus, a SARS-CoV-2 variant may be reclassified over time as: *a)* Formerly Monitored Variant (FMV); *b)* previously circulating VOI/VOC (ex-VOI/VOC); *c)* VOC Lineage Under Monitoring (VOC-LUM), with characteristics which might turn out to be substantially different from the main lineage.

In the present study, we fitted the WHO data [1, 2] by exploiting a function which exclusively depends on the number of infected cases worldwide. Our fit can predict an estimate of the number of new relevant variants per ten million cases and, consequently, the cumulative number of relevant variants in a given epidemiological situation.

As shown in this research, the number of new relevant variants per ten million cases decreases very slowly as the cumulative number of Covid-19 cases increases. Therefore, it becomes crucial to carefully monitor and reduce virus circulation in order to avoid the emergence of new variants, which may not be suitably covered by the vaccines and drugs currently available [8].

Although it is obvious that the number of relevant variants increases with the cases throughout the world, it is extremely difficult to find out the precise relationship between these two variables. Our model is a simple attempt to make a fairly reliable estimate of the risk of new variants that can impact public health as the virus continues to spread.

## Methods

By means of Wolfram Mathematica [9] we fitted the data on SARS-CoV-2 variants in order to evaluate the cumulative number *v* of relevant SARS-CoV-2 variants versus the cumulative number *N* of infected subjects worldwide.

The function *v* fitting the WHO data must satisfy the following conditions:

1. If there is no infection, the number of variants is zero; vice versa, if the virus replicates infinite times, the cumulative number of variants is also infinite.
2. The cumulative number of variants increases with the number of infections, i.e. as the virus replications increase.
3. As the cumulative number of variants increases with the total cases in the world, the emergence of new virus mutations turns out to be slightly less frequent.

The fit of WHO data was obtained by means of the following function:

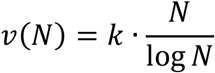

where *k* is the constant of the numerical fit and “log *N*” represents the natural logarithm of *N*. This function satisfies all the previous conditions, as shown in Appendix A.

The choice of the function *v* = *k* · *N*/ log *N* exploited in the fit can be justified through the heuristic arguments discussed in Appendix B, concerning both the biology of virus replication and well-known results from number theory and physics.

In Appendix C we fitted the WHO data by using another class of functions which satisfy the conditions listed in this section. In Appendix D we performed the fit discussed here by exploiting the ECDC data on relevant SARS-CoV-2 variants [6, 7] instead of using the WHO data [1, 2]. Finally, Appendix E reports further details on the numerical fit and the approximation given by linear regression.

## Results

We fitted the data recorded by WHO [1, 2] up to March 2022 by means of a specific code written with Wolfram Mathematica 13.0.1 [9].

Table 1 lists the characteristics of SARS-CoV-2 variants reported by WHO [1, 2]: date and country of the earliest detection, PANGO (Phylogenetic Assignment of Named Global Outbreak) and WHO classification, current relevance, total number of cases in the world at the end of the month of detection and cumulative number of variants. PANGO is a rule-based nomenclature system for naming and tracking SARS-CoV-2 genetic lineages [10].

**Table 1.**
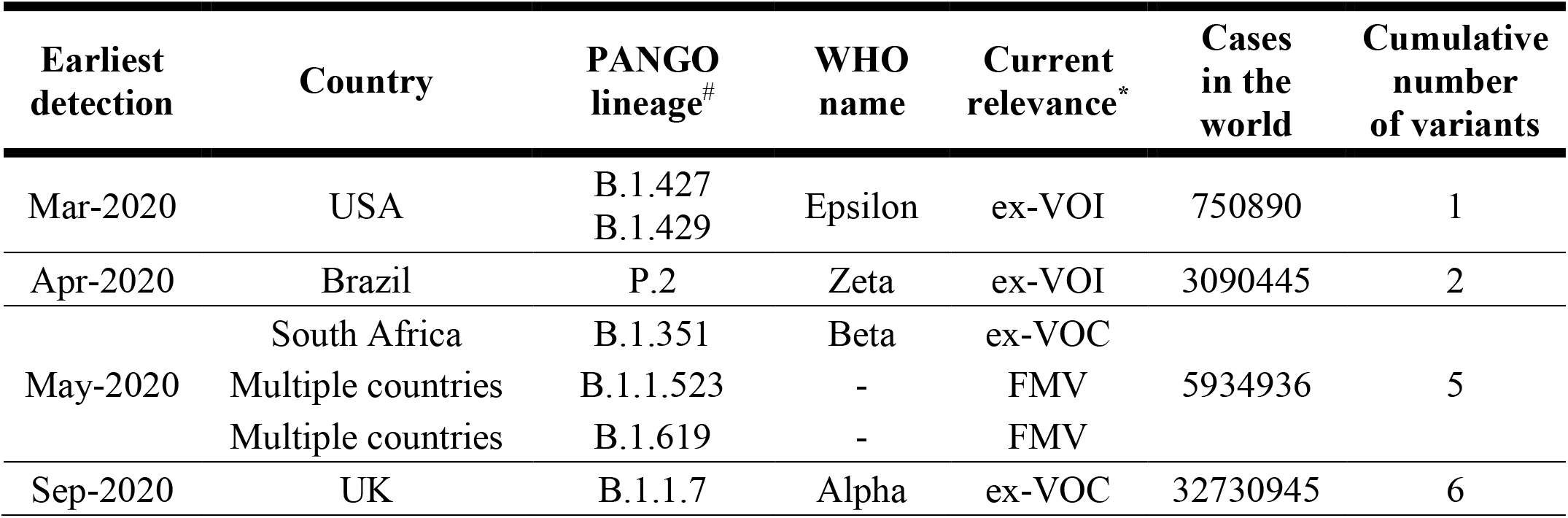
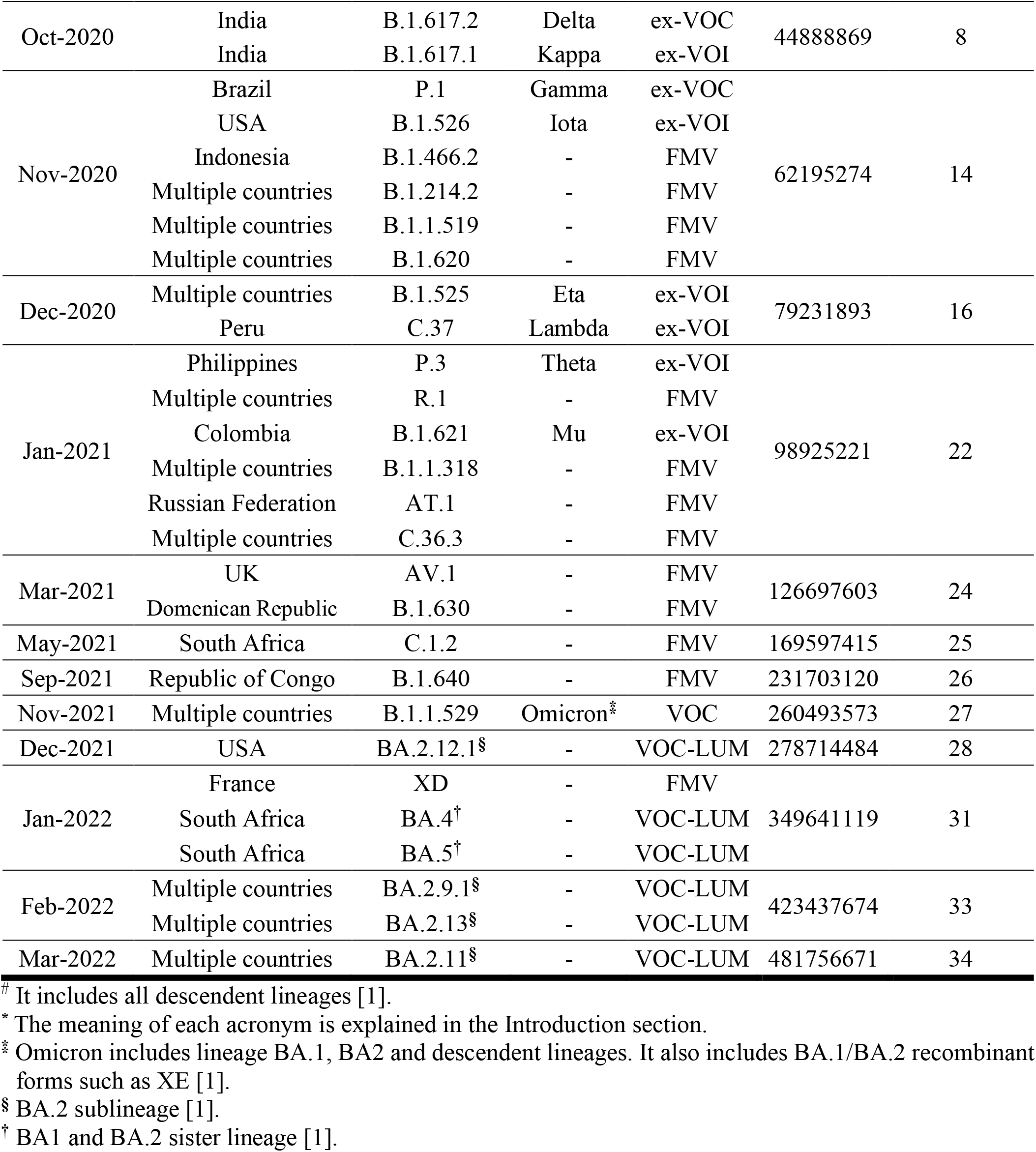
Characteristics of SARS-CoV-2 variants recorded by WHO [1, 2]: date and country of the earliest detection, PANGO and WHO nomenclature, current relevance and cumulative number of cases in the world at the end of the month of detection. The last column summarises the cumulative number of observed relevant variants.

The numerical fit of the WHO data was obtained by means of the function *v* (*N*) = *k* ·*N*/ log *N*, where the constant of the numerical fit is *k* = 1.83 · 10^−6^. The 95% confidence interval (CI) of *k* is given by 95% CI = (1.52 – 2.15) · 10^−6^. The adjusted *R*-squared, measuring the goodness of the fit, turned out to be *R*^2^ = 0.91.

Figure 1 represents the cumulative number *v* of relevant SARS-CoV-2 variants versus the cumulative number *N* of cases in the world. The dots from 1 to 16 correspond to the data reported by WHO [1, 2] from March 2020 to March 2022; the solid line represents the function *v* = *k* · *N*/ log *N* used in the fit with Wolfram Mathematica.

**Figure 1.**
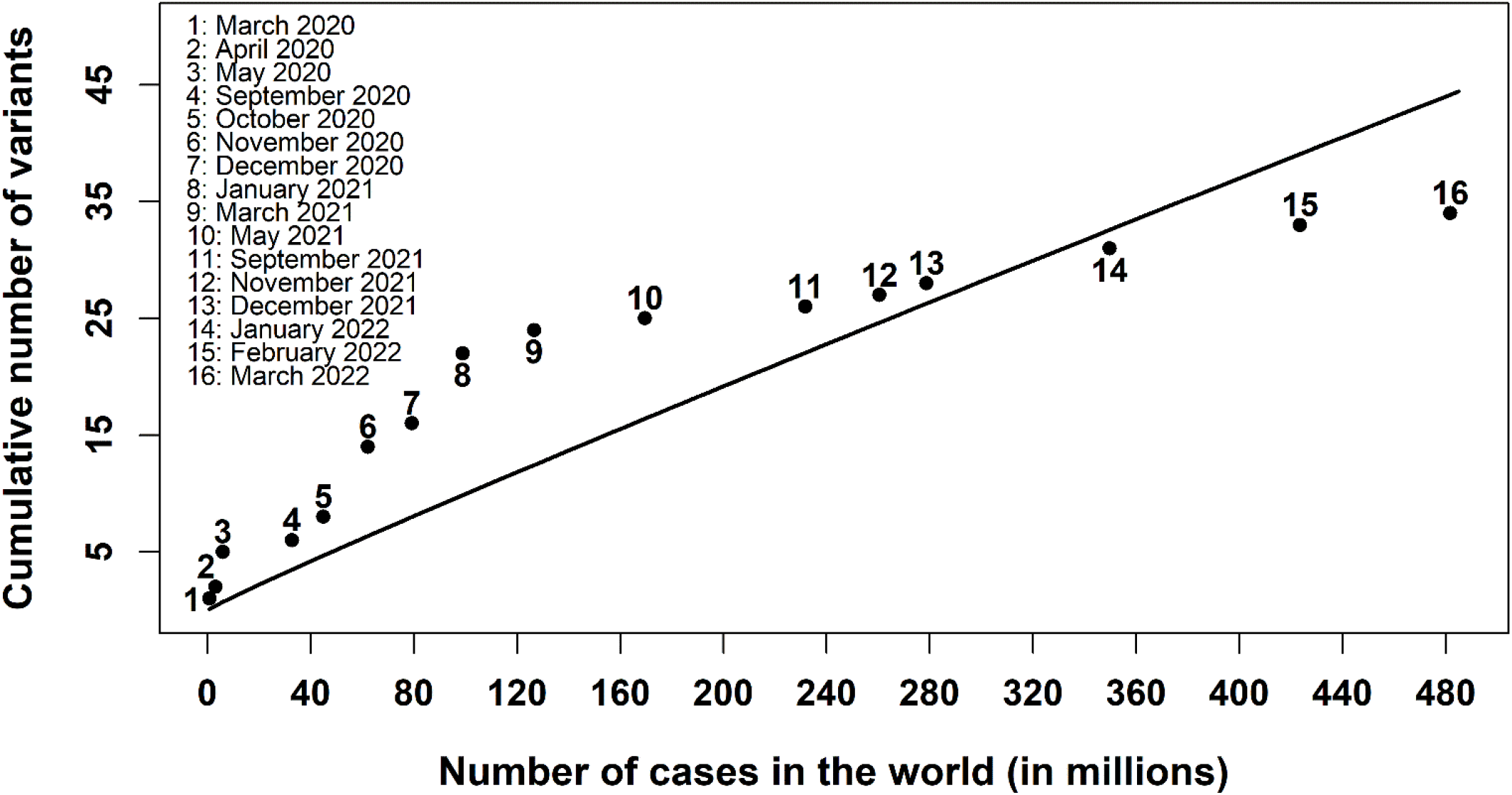
Cumulative number of relevant SARS-CoV-2 variants versus the cumulative number of cases in the world. The dots from 1 to 16 indicate the data reported by WHO [1, 2] from March 2020 to March 2022; the solid line represents the function *v* = *k* · *N*/ log *N* used in the numerical fit with Wolfram Mathematica.

In Appendix E we discussed the error associated to our estimates; for instance, in January 2022 the evaluation through our model of the number of relevant SARS-CoV-2 variants and the corresponding 95% CI turned out to be 32.6 (27.0 – 38.2), while the WHO value was 31. The residual, given by the difference between observed and predicted values, was 1.6.

The residuals and the 95% CIs of our estimates, listed in Appendix E, represent the uncertainty associated to the goodness of our mathematical fit but exclude the error related to the data provided by WHO for infected cases in the world and relevant variants, which were probably underestimated. The maximum absolute value *r*_*max*_ of the residuals was *r*_*max*_ = 12.15, corresponding to the 8^th^ observation in Figure 1; the residual standard deviation was *σ*_*r*_ = 6.75.

As discussed in Appendix A, the number *n* of new relevant variants per ten million (10^7^) cases is given by 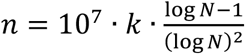 which becomes 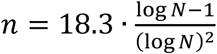 by substituting the numerical value of *k*. Figure 2 represents the number *n* of new relevant SARS-CoV-2 variants per ten million cases versus the cumulative number *N* of cases in the world.

**Figure 2.**
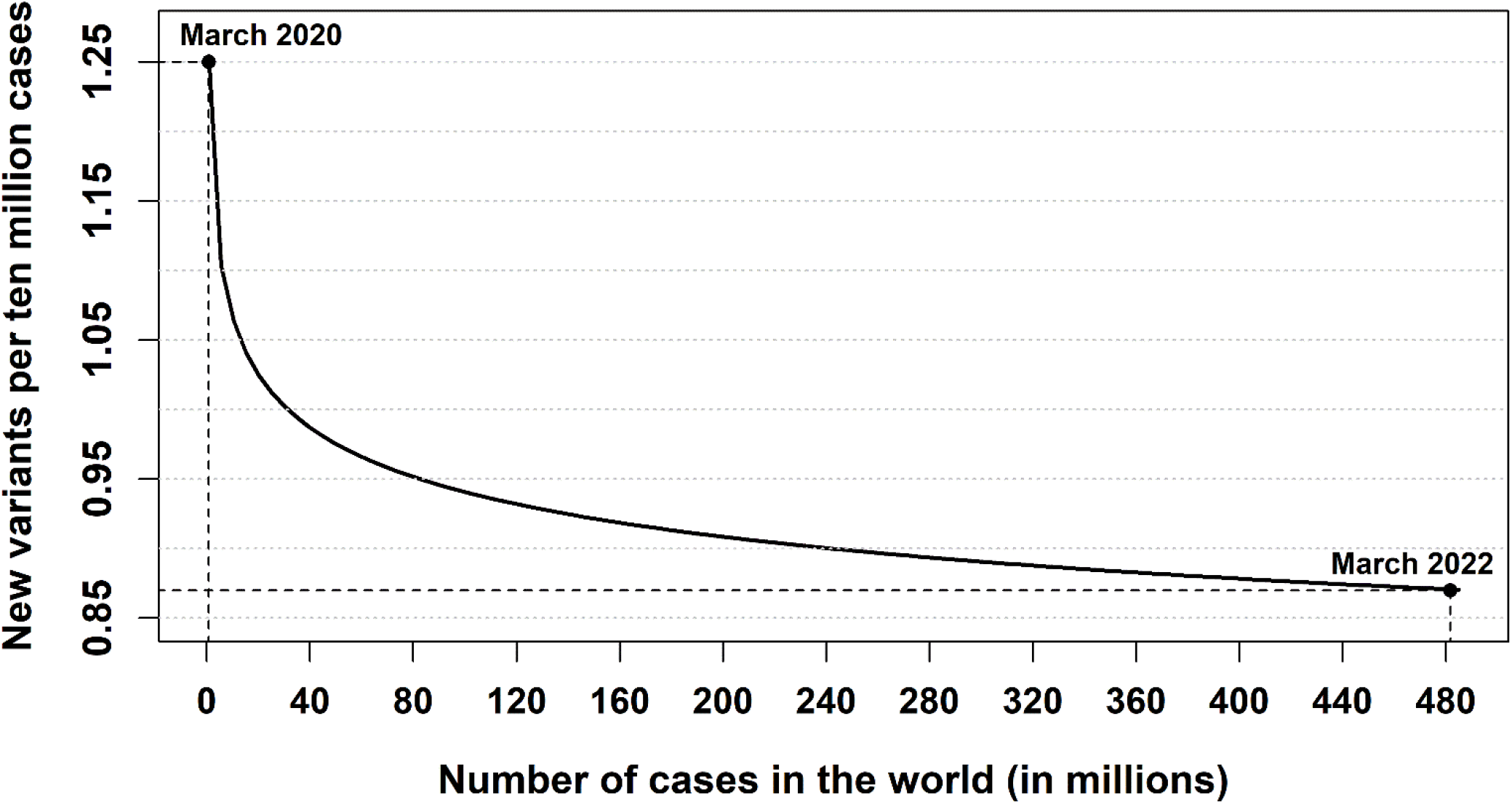
Number *n* of new relevant SARS-CoV-2 variants per ten million cases versus the cumulative number of cases in the world. From March 2020 to March 2022 *n* decreased from 1.25 to 0.87.

As shown in Appendix A, the relative variation |*n*^′^(*N*)| of the number *n* of new relevant variants per ten million cases is given by 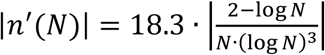, therefore it turns out |*n*^′^(*N*)| ≪ 1 for *N* ≫ 1. This analytical result implies that the number *n* of new relevant variants per ten million cases decreases very slowly as the virus continues to circulate. For instance, between March 2020 and March 2022 *n* decreased by 30.4%, from 1.25 to 0.87. However, a further reduction by 10%, from 0.87 to 0.78, would require that the cumulative cases in the world increase to 5.6 billion, i.e. ten times the total cases from the beginning of the epidemic up to June 2022.

Appendix E reports the values of the parameter *n*, with the corresponding 95% CIs, for all of the sixteen WHO observations; for instance, in March 2020 and March 2022 we had *n* = 1.25 (1.04 − 1.47) and *n* = 0.87 (0.72 − 1.02), respectively.

The relative standard error on the values *v* and *n* predicted by the model is (*k*_2_ − *k*_1_)/ (2*t*′ · *k*) = 8.1%, where *k*_1_ = 1.52 · 10^−6^ and *k*_2_ = 2.15 · 10^−6^ are the lower and upper limits of the 95% CI of the parameter *k* = 1.83 · 10^−6^ in the fit function and *t*’ = 2.13 is the two-sided 5% point of the Student’s t distribution with 15 degrees of freedom [11].

Our model cannot predict the *exact* number of relevant variants but only an estimate, since the errors associated to both the mathematical fit and the WHO data are not negligible. For instance, our fit shows that, up to June 2022, the cumulative number of relevant SARS-CoV-2 variants has reached the order of magnitude of few tens. According to our model, the number of cumulative relevant variants will become 100 for 1.14 billion infected cases, corresponding to about twice the total infections in the world from the beginning of the epidemic up to June 2022.

Figure 3 represents the predicted increase of the cumulative number of relevant variants per each step of ten million infections, from 480 to 600 million cumulative cases in the world. In this range of cases, the number of relevant variants is predicted to increase from 44.0 to 54.4.

**Figure 3.**
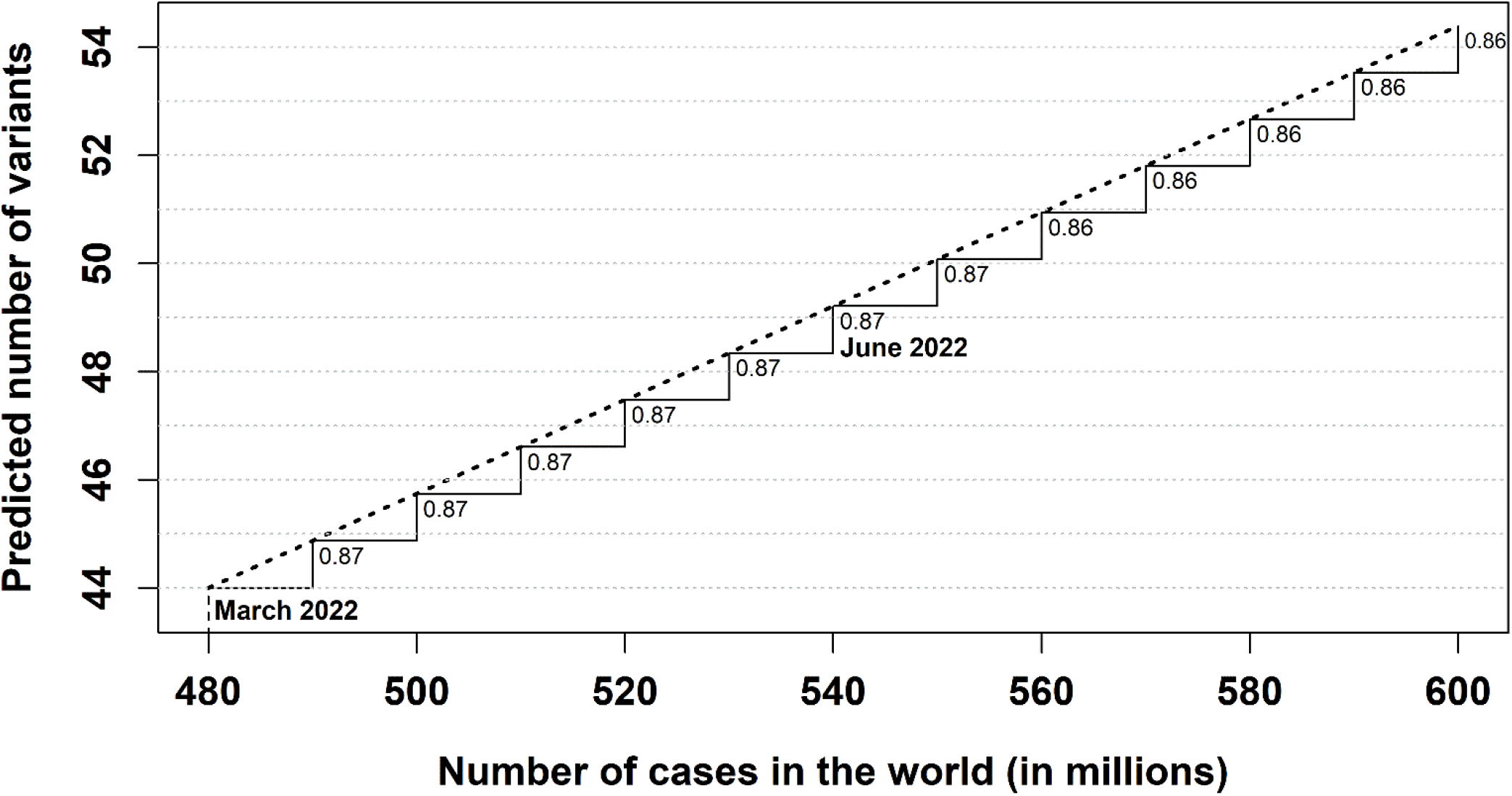
Prediction of the cumulative number of SARS-CoV-2 variants from 480 to 600 million cases in the world. The dotted line represents the function *v* = *k* · *N*/ log *N*, while each step shown in the plot corresponds to the predicted number of new relevant variants per ten million cases.

As reported in Figure 3, the total infected cases were about 480 million at the end of March 2022 and about 540 million at the end of June 2022.

Our model only focuses on the relationship between the number of virus replications and the emergence of relevant variants. All other factors involved in the diffusion of new variants were not taken into account. For this reason, we supposed that the parameter *k* in the fit *v* = *k* · *N*/ log *N* is constant, although it actually varies with all other factors affecting the emergence of relevant variants.

In Appendix D the fit discussed in this section by using the WHO data on SARS-CoV-2 variants [1, 2] was performed by exploiting the ECDC data [6, 7]. The value obtained for the constant *k* in the fit function turned out to be compatible with that derived from the WHO data, since the 99% CIs of *k* in the ECDC and WHO fits overlapped.

## Discussion

Since the start of the Covid-19 pandemic there has been an impressive global effort in investigating every aspect of coronavirus spread [12], including immunogenetic [13, 14] and epidemiological [15] issues.

In this study, we built a simple mathematical model to predict, from the number of infected cases in the world, both the cumulative number of relevant SARS-CoV-2 variants and the number of new relevant variants in a given epidemiological situation.

In particular, we found that the number of new relevant variants per ten million cases diminished by 30.4%, from 1.25 to 0.87, between March 2020 and March 2022. However, this decrease is getting slower as the virus continues to circulate, so that a further reduction by 10%, to about 0.78, will happen only if the total infected cases in the world reach about ten times the infections recorded from the start of the epidemic up to June 2022. Therefore, the persistence of virus circulation is bound to always cause the emergence of new relevant SARS-CoV-2 variants.

In Appendix B we justified the choice of the function exploited in the fit through arguments relied on the biological mechanism underlying virus replication or inspired to results derived from mathematics and physics. The shape of the function is fundamental for the goodness of the fit, therefore it was important to reinforce the arguments behind the choice of the analytical expression of the function.

Our method depends critically on the WHO efficiency in tracking the most relevant SARS-CoV-2 variants. For this reason, in Appendix C the fit was performed by exploiting the ECDC data on relevant SARS-CoV-2 variants instead of using the WHO data: the value of the fit parameter *k* obtained from the ECDC data turned out to be compatible with the value derived from WHO data.

The model discussed in this research for the SARS-CoV-2 variants can be extended to the evaluation of the number of relevant variants of any other virus if accurate data on those variants are available.

Our model does not provide the *exact* number of relevant SARS-CoV-2 variants but only an estimate of its order of magnitude, since the uncertainty of the mathematical fit and the errors associated to WHO data are not negligible. In Appendix E we reported the residuals and the 95% CIs of our estimates, corresponding to a relative standard error on the predicted values which turned out to be about 8%.

The diffusion of new relevant variants depends on a large variety of factors. For instance, the ability to monitor all virus mutations, the effectiveness of containment measures and vaccination campaigns, the evolution patterns of the virus [16], and so on. All these factors were ignored in our model, which only focuses on the number of virus replications. The parameter *k* appearing in the fit *v* = *k* · *N*/ log *N* is not actually constant, as supposed in our model, but varies with all the other factors which can affect the emergence of relevant variants. None the less, our model provides a fairly good estimate of what we can expect in the future.

SARS-CoV-2 vaccination has led to a decrease in hospitalisations and disease severity. However, the current number of infections is still too high to prevent the appearance of new variants potentially dangerous to public health.

The number of variants increases with the number of cases *in the world*: this result underlines the urgency of making every effort to reduce the impact of virus replication in all geographical areas. The risk that new relevant variants may emerge anywhere in the world indicates that the winning strategy is not to leave any country behind in the battle against the virus.

The possibility to predict the number of new relevant SARS-CoV-2 variants will become increasingly important in future to ensure optimal planning of vaccination campaigns by healthcare services, united in the awareness that new variants can change the characteristics of the virus and greatly influence the global management of the pandemic.

## Data Availability

All data referred to in the manuscript are available online at https://www.who.int/en/activities/tracking-SARS-CoV-2-variants

## Authors’ contributions

The authors contributed equally to the article.

## Ethical approval

Not applicable.

## Acknowledgments

The authors are grateful to Anna Maria Koopmans for translations, professional writing assistance and preparation of the manuscript.

