## Appendices for "How many relevant SARS-CoV-2 variants might we expect in the future?"

### Appendix A

#### Mathematical details on the fit function

In this Appendix we specify some technical details which complete the discussion reported in the Methods section of the article.

The function exploited to fit the WHO data is

$$v(N) = k \cdot \frac{N}{\log N}$$

where $v$ is the cumulative number of relevant SARS-CoV-2 variants, $N$ is the cumulative number of infected subjects worldwide, $k$ is the constant of the numerical fit and "$\log N$" represents the natural logarithm of $N$.

This function satisfies all the conditions listed in the Methods section of the article, as shown below:

1) the function $v$ varies from zero to infinity:

$$\lim_{N \to 0} v = 0 \quad \text{and} \quad \lim_{N \to \infty} v = \infty$$

2) The function $v$ increases monotonically, therefore the first derivative $v'(N)$ is positive:

$$v'(N) = k \cdot \frac{\log N - 1}{(\log N)^2} > 0 \text{ for } N > e$$

where $e \cong 2.72$ is the Euler's number. In this research $N \gg 1$, hence $v'(N) > 0$.

3) The first derivative of $v$ decreases monotonically, therefore the second derivative $v''(N)$ is negative:

$$v''(N) = k \cdot \frac{2 - \log N}{N \cdot (\log N)^3} < 0 \text{ for } N > e^2$$

with $e^2 \cong 7.39$. In our study it always turns out $N \gg 1$, therefore $v''(N) < 0$.

The number $n$ of new relevant variants per ten million cases ($\Delta N = 10^7$) turns out to be:

$$n \cong v'(N) \cdot \Delta N = 10^7 \cdot k \cdot \frac{\log N - 1}{(\log N)^2}$$

The relative variation $|n'(N)|$ of new relevant variants per ten million cases decreases as the number $N$ of cases increases: $|n'(N)| \cong |v''(N)| \cdot \Delta N = 10^7 \cdot k \cdot \left|\frac{2-\log N}{N \cdot (\log N)^3}\right|$. Being $N \gg 1$, it turns out $|n'(N)| \ll 1$.

### Appendix B

#### Heuristic arguments on the fit function

In this Appendix we present two heuristic arguments supporting the choice of the function exploited in our study to fit the WHO data on relevant SARS-CoV-2 variants: the first argument is inspired to the model introduced by Delbrück and Luria on bacterial mutations, the second one relies on a possible relationship between virus variants and prime numbers.

##### DELBRÜCK AND LURIA MODEL

In 1943 Delbrück and Luria studied the effect of a virus on bacterial cultures [B1]. They found that bacterial variants resistant to the action of the virus appeared as a consequence of mutations which occurred independently of the virus (hypothesis of mutation to immunity) rather than induced by the virus (hypothesis of acquired hereditary immunity). We extended the same approach to the investigation of relevant mutations in a viral sample, including virus variants resistant to vaccines.

Following Delbrück and Luria [B1], the number $N_t$ of bacteria present at time $t$ in a growing culture is expressed by the equation

$$N_t = N_0 e^t$$

where $N_0$ is the initial number of bacteria and $t$ is measured in units of division cycles of the bacteria.

The likely average $r$ of the resistant bacteria in a limited number $C$ of samples is given by

$$r = aN_t \ln(N_t Ca)$$

where $a$ is the mutation rate, i.e. the chance of mutation per bacterium per time unit.

The variance of the distribution of all resistant bacteria in a limited number of $C$ cultures is

$$var_r = Ca^2 {N_t}^2$$

By comparing the variance $var_r$ with the likely average $r$ of resistant bacteria, Delbrück and Luria found:

$$var_r = r \cdot Ca \cdot \frac{N_t}{\ln(N_t Ca)}$$

Based on the hypothesis of random mutations, the number of resistant bacteria is not distributed according to Poisson's law and the variance $var_r$ turns out to be much greater than the likely average $r$ of resistant bacteria. On the contrary, the hypothesis of acquired hereditary immunity predicts that variance and average should be equal, according to Poisson's law. The experiment performed by Delbrück and Luria confirmed the hypothesis of random mutations [B1].

If we assume that the variance $var_r$ is proportional to both the likely average $r$ of resistant bacteria and the number $v$ of mutations, we can write: $var_r = b \cdot v \cdot r$, where $b$ is a constant of proportionality. By substituting this expression of $var_r$ in the equation connecting variance $var_r$ and likely average $r$, we get:

$$v = C\frac{a}{b} \cdot \frac{N_t}{\ln(N_t C a)}$$

If we define a new constant $k = C\frac{a}{b}$ and suppose $|\ln(Ca)| \ll \ln N_t$, the previous equation becomes

$$v = k \cdot \frac{N}{\log N}$$

with $N \equiv N_t$ and $\log \equiv \ln$.

The findings of Delbrück and Luria [B1] on bacterial mutations can be adapted to the study of virus mutations and thus allow the evaluation of the number of SARS-CoV-2 variants.

The equation $v = k \cdot \frac{N}{\log N}$ obtained from the Delbrück and Luria model can be interpreted as the number of relevant virus variants $v$ up to a time $t$ as function of the number $N$ of infections up to that time. Delbrück and Luria [B1] considered a bacterial culture growing according to an exponential law. Therefore, the equation obtained from their model is strictly valid only in the exponential phases of the Covid-19 pandemic. Moreover, Delbrück and Luria considered bacterial mutations resistant to a virus while the viral variants in our study are either mutations resistant to vaccines or virus variants with relevant characteristics concerning transmissibility, disease course and global public health.

#### VIRUS VARIANTS AND PRIME NUMBERS

The connection discussed here between virus mutations, quantum states and prime numbers yields a suggestive justification of the analytic form of the function used for the fit of WHO data, although a precise theoretical framework is still lacking.

**Prime numbers.** As discussed in the Methods section, the fit of WHO data was obtained by means of the function $v(N) = k \cdot N/\log N$, where $k$ is the constant of the numerical fit.

In 1801 Gauss [B2] found that the function $\pi(x) \sim \frac{x}{\log x}$ yields asymptotically (i.e. for $x$ sufficiently large) the cumulative number of primes less than a given number $x$. By comparing the Gauss function $\pi(x)$ and the function exploited for the fit, it is clear that the cumulative number of relevant variants $v$ for $N$ total infected subjects in the world is proportional to the cumulative number $\pi(N)$ of primes less than $N$, i.e. $v(N) = k \cdot \pi(N)$.

A more accurate expression of the number of primes less than $x$ is given by the logarithmic integral function $Li(x)$, defined as $Li(x) = \int_0^x \frac{dt}{\log t}$ . By exploiting the logarithmic integral function for the fit of WHO data, the cumulative number $\hat{v}$ of relevant SARS-CoV-2 variants for $N$ infected cases in the world becomes: $\hat{v}(N) = h \cdot Li(N) = h \cdot \int_0^N \frac{dt}{\log t}$ with the constant $h$ given by $h = 1.73 \cdot 10^{-6}$ and $95\%\ \mathrm{CI} = (1.44 - 2.03) \cdot 10^{-6}$. The adjusted $R$-squared, measuring the goodness of the fit, is $R^2 = 0.91$ with both the Gauss function and the logarithmic integral function.

The difference between the fits $v(N)$ and $\hat{v}(N)$ is less than 1 for $N \leq 2.32 \cdot 10^9$, therefore in the current range of $N$ values we can use the simpler function $v(N)$.

The Gauss function provides an asymptotic approximation of the number of primes less than a sufficiently large quantity, therefore we considered the number of infected subjects all over the world instead of focusing on a specific country or geographical area, where the infections are a fraction of the world cases.

**Zeta function and quantum states.** In 1859 Riemann [B3] found an exact expression for the number of primes less than a given quantity. Riemann's formula involves a sum over the zeroes of the so-called zeta function $\zeta(s)$, whose relevant zeroes have all real part $\frac{1}{2}$ according to Riemann's Hypothesis [B4]. The distribution of the zeroes of $\zeta(s)$ along the critical line $z = \frac{1}{2}$ determines the distribution of the prime numbers, as well established in number theory [B5].

In 1977 Montgomery [B6] found the function which describes the spacing between the zeroes of $\zeta(s)$. As spotted by the physicist Dyson, such function is the same as that describing the spacing between the energy levels of heavy atomic nucleus. The link between zeroes of $\zeta(s)$ and quantum states also extends to chaotic systems, as pointed out by Berry [B7] and confirmed numerically by Odlyzko [B8].

In conclusion, a connection seems to exist between the distribution of the prime numbers and the quantum states of a physical system.

**Virus mutations.** In 1944 Schrödinger [B9], in his renowned essay "What is life?", suggested - following Delbrück [B10] - that a genetic mutation can be considered a sort of "quantum jump", i.e. a transition between two different states of a quantum system. Analogously, a virus variant may be interpreted as a genetic mutation due to a quantum transition between two different configurations in the structure of the virus.

The following figure schematises the connection between the cumulative number of primes less than a given quantity and the number of relevant variants for a given number of cases. The intermediate links in the scheme are the distribution of the zeroes of Riemann's zeta function and the spacing between the quantum states of a physical system.

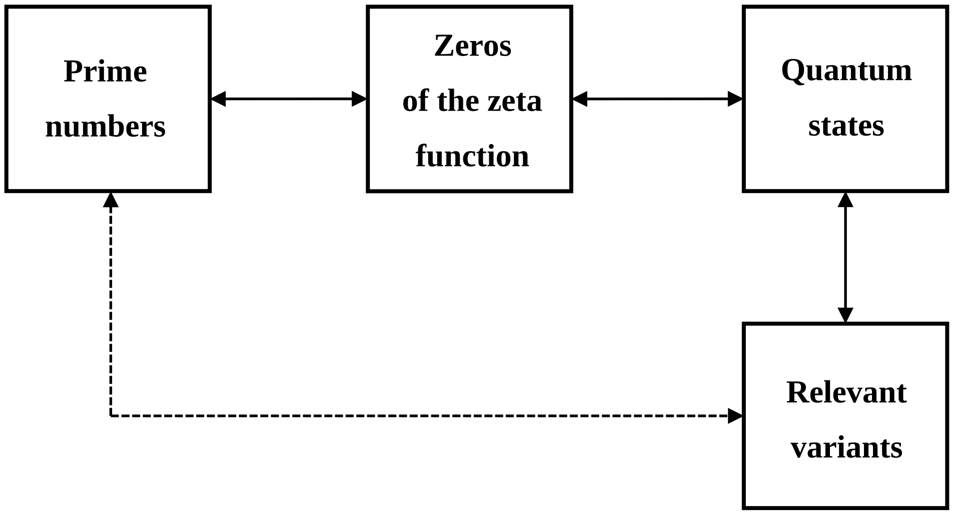

The numerical fit of WHO data could be performed by exploiting a great variety of functions different from the one we used (see Appendix C). However, our choice is supported by the suggestive hypothesis of a connection between virus variants and prime numbers inspired by quantum physics and number theory.

[B2] Gauss CF. Disquisitiones Arithmeticae. Leipzig: Fleischer; 1801.

[B3] Riemann B. On the Number of Prime Numbers less than a Given Quantity. Berlin: Monatsberichte der Berliner Akademie; 1859.

[B4] Edwards HM. Riemann’s Zeta Function. Mineola, New York: Dover Publications, Inc; 1974.

[B5] Hardy GH, Littlewood JE. Contributions to the theory of the Riemann zeta-function and the theory of the distribution of primes. Acta Math 1916; 41: 119-96. https://doi.org/10.1007/BF02422942.

[B6] Montgomery HL. Extreme values of the Riemann zeta function. Comm. Math Helv 1977; 52:511-18.

[B7] Berry MV. Riemann’s Zeta function: A model for quantum chaos? In: Seligman TH, Nishioka H (editors). Quantum Chaos and Statistical Nuclear Physics. Lecture Notes in Physics, vol 263. Berlin, Heidelberg: Springer; 1986. https://doi.org/10.1007/3-540-17171-1_1.

[B8] Odlyzko AM. On the distribution of spacings between zeroes of the zeta function. Mathematics of Computation 1987; 48 (177): 273-308. https://doi.org/10.2307/2007890.

[B9] Schrödinger E. What is Life? The Physical Aspects of the Living Cell. Cambridge: Cambridge University Press; 1944.

[B10] Timoféeff-Ressovsky NW, Zimmer KG, Delbrück M. Über die Natur der Genmutation und der Genstruktur. *Nachr. Ges. Wiss. Göttingen* 1935; Biologie: 189-245.

### Appendix C

#### Other fit functions

In order to fit the WHO data on relevant SARS-CoV-2 variants, we can use many functions satisfying the conditions discussed in the Methods section and in Appendix A, i.e. functions varying from zero to infinity, monotonically increasing and with the first derivative monotonically decreasing.

The functions $f = \alpha \cdot N^{\beta}$, with $\alpha > 0$ and $0 < \beta < 1$, satisfy all the requested conditions:

1) $\lim_{N \to 0} f = 0$ and $\lim_{N \to \infty} f = \infty$.

2) $f'(N) = \alpha \cdot \beta \cdot N^{-(1-\beta)} > 0$.

3) $f''(N) = -\alpha \cdot \beta \cdot (1-\beta) \cdot N^{-(2-\beta)} < 0$.

If we choose the value $\beta = \frac{1}{2}$, we obtain the square root function $f = \alpha \cdot \sqrt{N}$. In this case the fit of WHO data through Wolfram Mathematica yields the value $\alpha = 1.69 \cdot 10^{-3}$, with 95% CI = $(1.58 - 1.80) \cdot 10^{-3}$. The adjusted $R$-squared measuring the goodness of the fit is $R^2 = 0.98$, the maximum residual is $r_{max} = 5.20$ and the residual standard deviation is $\sigma_r = 2.60$ (see Appendix E for the definition of $r_{max}$ and $\sigma_r$).

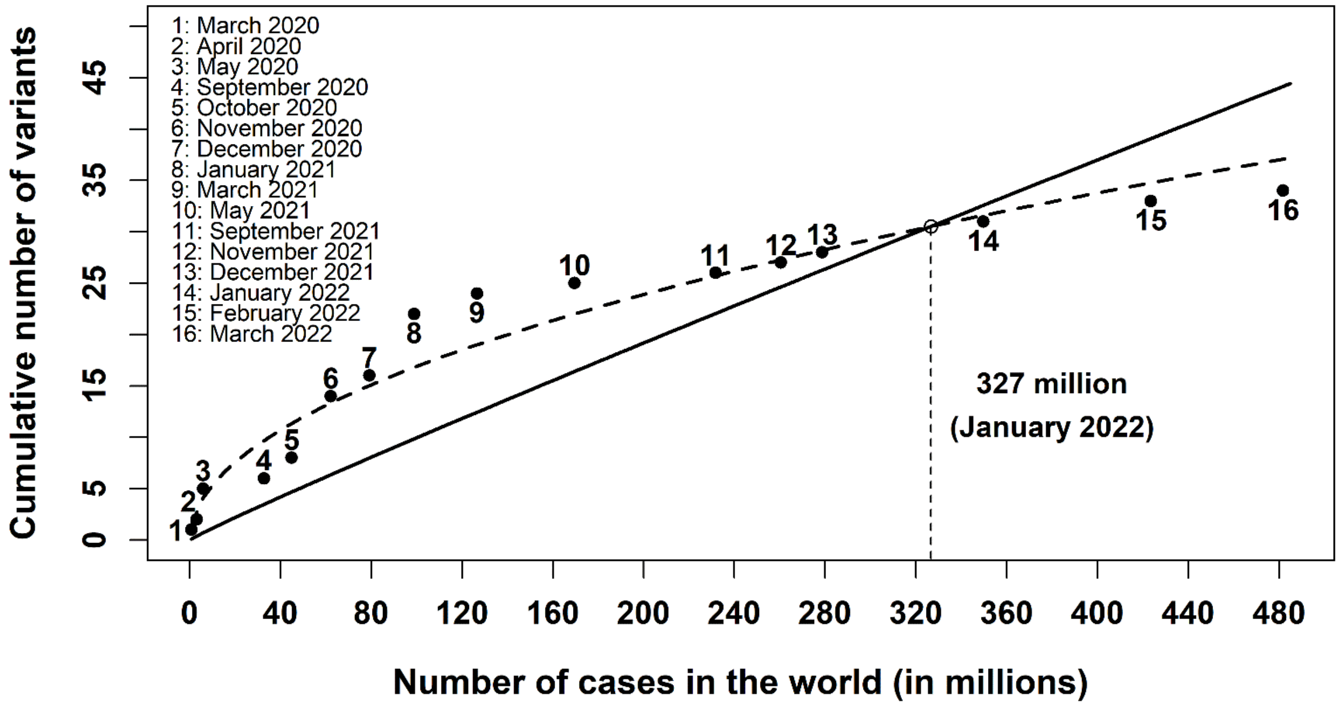

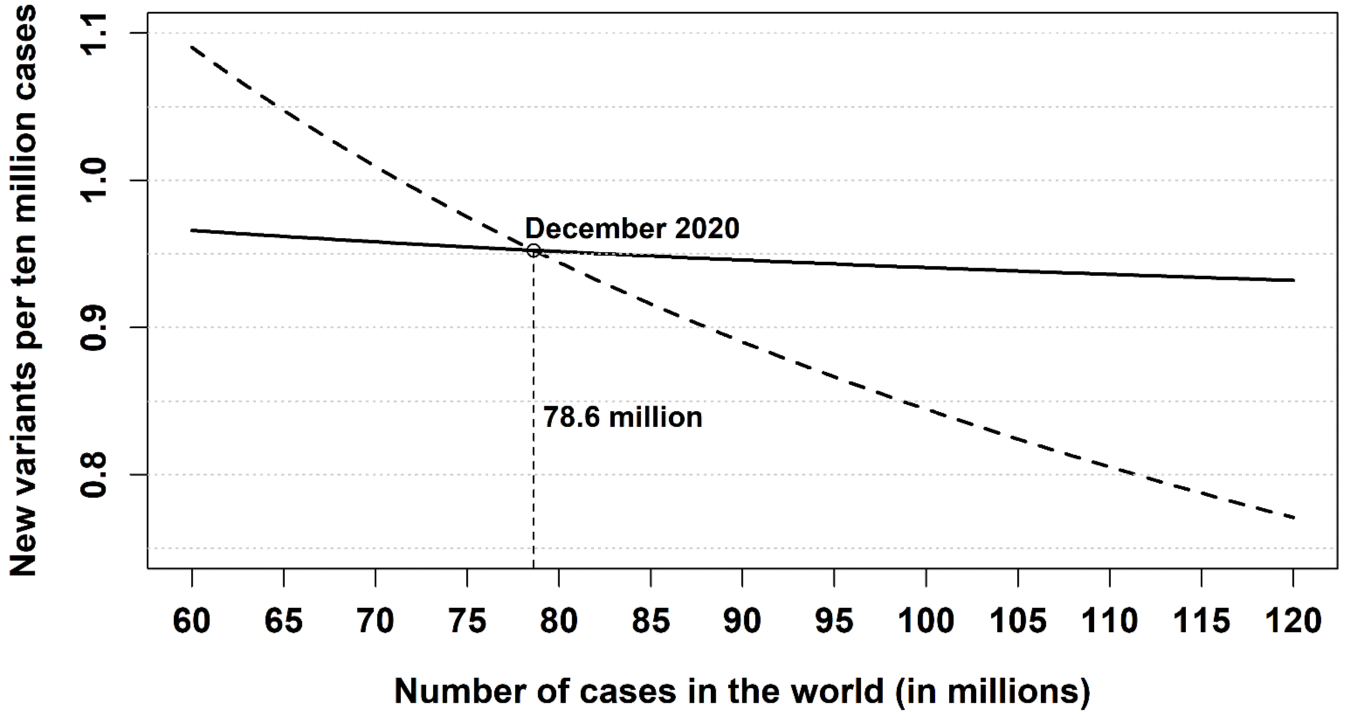


The previous figures represent the fits of WHO data through the logarithmic function $v = k \cdot N/\log N$, with $k = 1.83 \cdot 10^{-6}$ (solid line), and the square root function $f = \alpha \cdot \sqrt{N}$, with $\alpha = 1.69 \cdot 10^{-3}$ (dashed line). The first figure shows the cumulative number of relevant variants while the second figure represents the number of new relevant variants per ten million infected cases. The empty circle in each figure indicates the intersection of the two fits, i.e. their common value with the corresponding number of infected cases and the date of detection.

The number $n$ of new relevant variants per ten million cases is $n = \frac{k \cdot 10^7 (\log N - 1)}{(\log N)^2}$, with $k = 1.83 \cdot 10^{-6}$, for the logarithmic function (solid line) and $n = \frac{\alpha \cdot 10^7}{2\sqrt{N}}$, with $\alpha = 1.69 \cdot 10^{-3}$, for the square root function (dashed line).

As shown in the second figure, the number $n$ of new relevant invariants decreases much faster in the square root model than in the logarithmic fit. For instance, between January 2021 and January 2022 $n$ decreased by 46.8% (from 0.85 to 0.45) in the square root fit, while it only decreased by 6.4% (from 0.94 to 0.88) in the logarithmic model. The reduction between January 2022 and June 2022 was about 14.8% in the square root fit and only 2.0% in the logarithmic model.

The relative standard error on both the cumulative number of relevant variants predicted by the square root model and the number of new variants per ten million cases is given by $(k_2 - k_1)/(2t' \cdot k) = 3.0\%$, where $k_1 = 1.80 \cdot 10^{-3}$ and $k_2 = 1.58 \cdot 10^{-3}$ are the lower and upper limits of the 95% CI of the parameter $k = 1.69 \cdot 10^{-3}$ in the fit function and $t' = 2.13$ is the two-sided 5% point of the Student's t distribution with 15 degrees of freedom.

If we fit the WHO data with the function $f = \alpha \cdot N^{\beta}$, the best fit values for the parameters $\alpha$ and $\beta$ obtained with Wolfram Mathematica are $\alpha = 4.01 \cdot 10^{-3}$ and $\beta = 0.46 \cong \frac{1}{2}$.

The choice of the function $f = \alpha \cdot \sqrt{N}$ can be justified heuristically by considering the virus variants as "errors" in the virus replication process. Since the relative error affecting a measurement in a system with $N$ elements is proportional to $\frac{1}{\sqrt{N}}$ and the virus replications are proportional to the number $N$ of infected subjects, the absolute error on the virus replication – and hence the number of virus variants – turns out to be proportional to $N \cdot \frac{1}{\sqrt{N}} = \sqrt{N}$.

### Appendix D

#### Numerical fit from the ECDC data

In this Appendix we exploit the ECDC data on relevant SARS-CoV-2 variants instead of the WHO data. The parameter $\boldsymbol{k}$ in the fit function depends critically on the epidemiological data, therefore it is important to check if the value of $\boldsymbol{k}$ obtained through the ECDC data is compatible with the value derived from the WHO data in the Results section of the article.

The classification of SARS-CoV-2 variants reported by ECDC [D1] is presented in the box below.

| | |
|---|---|
| VARIANTS UNDER MONITORING (VUM)<br>There is some indication that these variants could have an impact on the epidemiological situation, but the evidence is weak or has not yet been assessed. | VARIANTS OF CONCERN (VOC)<br>Clear evidence is available indicating a significant impact of these variants on transmissibility and severity of SARS-CoV-2 and/or immunity, with critical effects on the epidemiological situation. |
| VARIANTS OF INTEREST (VOI)<br>Evidence is available that these variants could affect transmissibility, severity and/or immunity, realistically having an impact on the epidemiological situation. However, the evidence is still preliminary. | DE-ESCALATED VARIANTS<br>Variants which satisfy at least one the following criteria: 1) no longer circulating, 2) still circulating but without any impact on the overall epidemiological situation, 3) not associated with any concerning properties. |

Table D1 lists the characteristics of SARS-CoV-2 variants reported by ECDC [D1, D2]: date and country of the first detection, lineage and WHO classification, total number of cases in the world at the end of the month of detection and cumulative number of variants.

The year and month of the first detection of BA.2+L452X were not available (NA), therefore this variant has not been included in our fit, which requires to know the number of infected cases up to the date of the first detection.

**Table D1**. Characteristics of SARS-CoV-2 variants recorded by ECDC [D1, D2]: date and country of the first detection, lineage and additional mutations, WHO nomenclature, cumulative number of cases in the world at the end of the month of detection. The last column summarises the cumulative number of the relevant variants recorded by ECDC.

| Year and month first detected | Country first detected | Lineage[#] and additional mutations | WHO label[§] | Cases in the world[†] | Cumulative number of variants |
|---|---|---|---|---|---|
| Sep-2020 | United Kingdom | B.1.1.7 | Alpha | 33735630 | 3 |
| | USA | B.1.427/B.1.429 | Epsilon | | |
| | South Africa | B.1.351 | Beta | | |
| Oct-2020 | Unclear[*] | C.16 | - | 44085676 | 5 |
| | USA | B.1.526.1 | - | | |
| Nov-2020 | Mexico | B.1.1.519 | - | 63781233 | 6 |
| Dec-2020 | United Kingdom | B.1.1.7+E484K | - | 81242993 | 20 |
| | Nigeria | B.1.525 | Eta | | |
| | India | B.1.617.1 | Kappa | | |
| | Unclear[*] | B.1214.2 | - | | |
| | United Kingdom | A.23.1+E484K | - | | |
| | Unclear[*] | A.27 | - | | |
| | Unclear[*] | A.28 | - | | |
| | South Africa | B.1.351+P384L | - | | |
| | USA | B.1.526 | Iota | | |
| | USA | B.1.526.2 | - | | |
| | Egypt | C.36+L452R | - | | |
| | Peru | C.37 | Lambda | | |
| | Brazil | P.1 | Gamma | | |
| | India | B.1.617.2 | Delta | | |
| Jan-2021 | The Philippines | P.3 | Theta | 103568588 | 28 |
| | Unclear[*] | B.1.351+E516Q | - | | |
| | United Kingdom | B.1.1.7+L452R | - | | |
| | United Kingdom | B.1.1.7+S494P | - | | |
| | Brazil | P.2 | Zeta | | |
| | Russian Federation | AT.1 | - | | |
| | Colombia | B.1.621 | Mu | | |
| | Unclear[*] | B.1.1.318 | - | | |
| Feb-2021 | France | B.1.616 | - | 114489076 | 32 |
| | Unclear[*] | B.1620 | - | | |
| | India | B.1.617.3 | - | | |
| | Italy | P.1+P681H | - | | |
| Mar-2021 | United Kingdom | AV.1 | - | 127654472 | 33 |
| Apr-2021 | India | B.1.617.2+E484X | - | 147380497 | 36 |
| | India | B.1.617.2+Q613H | - | | |
| | India | B.1.617.2+Q677H | - | | |

| | | | | | |
|---|---|---|---|---|---|
| Jun-2021 | United Kingdom | AY.4.2 | - | 181744185 | 39 |
| | United Kingdom | B.1.617.2+K417N | - | | |
| | South Africa | C.1.2 | - | | |
| Sep-2021 | Congo | B.1.640 | - | 232671700 | 40 |
| Nov-2021 | South Africa/Botswana | BA.1 (VOC) | Omicron | 262417085 | 43 |
| | South Africa | BA.2 (VOC) | Omicron | | |
| | South Africa | BA.3 (VUM) | Omicron | | |
| Jan-2022 | South Africa | BA.4 (VOC) | Omicron | 378132775 | 46 |
| | United Kingdom | XF | - | | |
| | France | XD | - | | |
| Feb-2022 | South Africa | BA.5 (VOC) | Omicron | 436615887 | 47 |
| NA | NA | BA.2+L452X (VOI) | - | NA | 48 |

### All sub-lineages of the listed lineages are included in the variant, e.g. BA.1.1 is included in Omicron BA.1 as it is a sub-lineage of BA.1 [D1].

* The earliest detections from several different countries are close in time and there is no clearly demonstrated travel link to a specific country that explains the detections [D1].

† The number of cumulative cases was obtained from Ref. [D2].

§ WHO labels are reported in Ref. [D3].

The numerical fit of the ECDC data was obtained by means of the function $v(N) = k \cdot N/\log N$, where the constant of the numerical fit is $k = 2.92 \cdot 10^{-6}$. The 95% confidence interval (CI) of $k$ is given by 95% CI $= (2.34 - 3.50) \cdot 10^{-6}$. The adjusted $R$-squared, measuring the goodness of the fit, turned out to be $R^2 = 0.90$.

The result obtained from the ECDC data is compatible with that obtained from the WHO data, as shown by the fact that the 99% CIs of the $k$ parameter in the two fits overlap:

$$99\%\ \text{CI (ECDC)} = (2.09 - 3.74) \cdot 10^{-6} \quad vs \quad 99\%\ \text{CI (WHO)} = (1.40 - 2.27) \cdot 10^{-6}$$

Figure D1 represents the cumulative number $v$ of relevant SARS-CoV-2 variants versus the cumulative number $N$ of cases in the world. The dots from 1 to 13 correspond to the data reported by ECDC [D1, D2] from September 2020 to February 2022; the solid line represents the function $v = k \cdot N/\log N$ used in the fit with Wolfram Mathematica.

As discussed in Appendix E for WHO data, in the fit of ECDC data the maximum absolute value $r_{max}$ of the residuals (differences between observed and predicted values) is $r_{max} = 16.99$, corresponding to the last observation in Figure D1. The residual standard deviation is $\sigma_r = 10.41$.

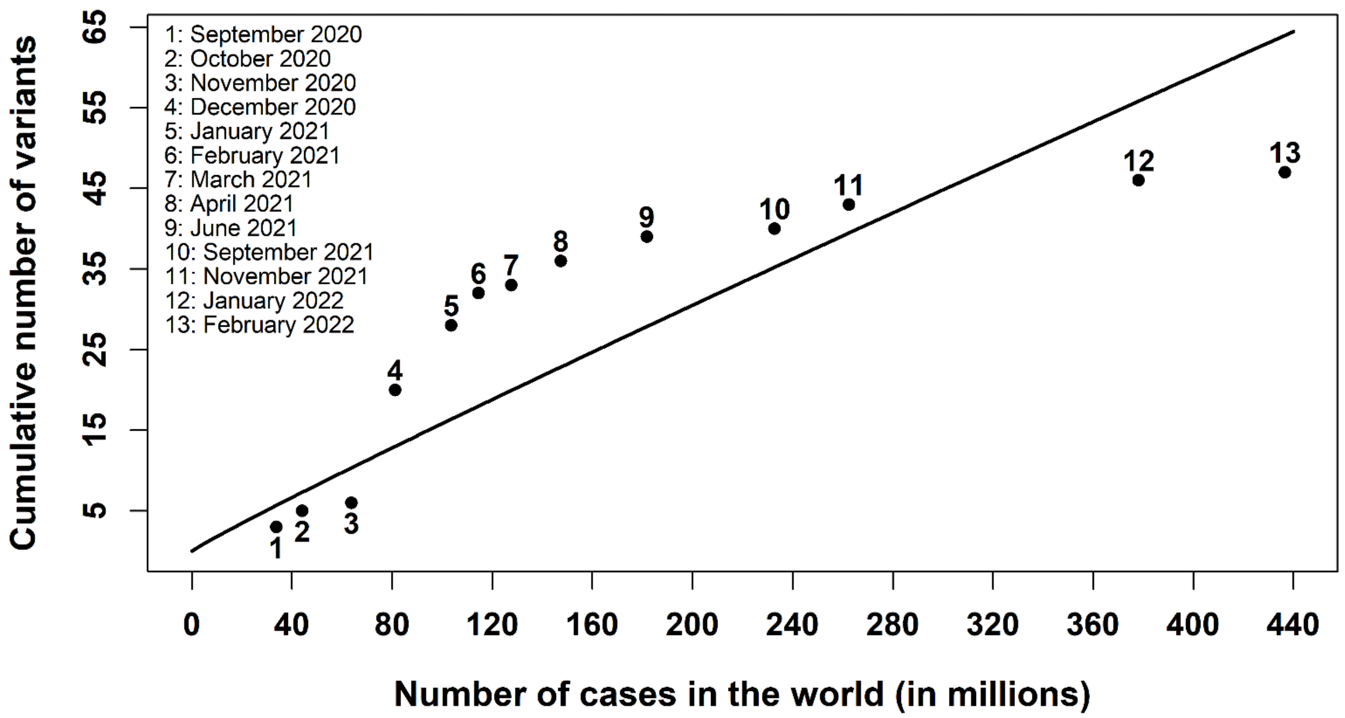


**Figure D1**. Cumulative number of relevant SARS-CoV-2 variants versus the cumulative number of cases in the world. The dots from 1 to 13 indicate the data reported by ECDC [D1, D2] from September 2020 to February 2022; the solid line represents the function $v = k \cdot N/\log N$ used in the numerical fit with Wolfram Mathematica.

The number $n$ of new relevant variants per ten million ($10^7$) cases is $n = 10^7 \cdot k \cdot \frac{\log N - 1}{(\log N)^2}$, which becomes $n = 29.2 \cdot \frac{\log N - 1}{(\log N)^2}$ by substituting the numerical value of $k$.

From September 2020 to February 2022 the number of new variants per ten million cases decreased by 12.6%, from 1.59 to 1.39. A further reduction by 10%, from 1.39 to 1.25, would require that the cumulative cases in the world increase to 4.7 billion, i.e. ten times the total cases from the beginning of the epidemic up to February 2022. This result shows that the number $n$ of new relevant variants per ten million cases decreases very slowly as the virus continues to circulate.

The relative standard error on the values $v$ and $n$ predicted by the model is $(k_2 - k_1)/(2t' \cdot k) = 9.2\%$, where $k_1 = 2.34 \cdot 10^{-6}$ and $k_2 = 3.50 \cdot 10^{-6}$ are the lower and upper limits of the 95% CI of the parameter $k = 2.92 \cdot 10^{-6}$ in the fit function and $t' = 2.18$ is the two-sided 5% point of the Student's t distribution with 12 degrees of freedom.

Figure D2 reports the number $n$ of new relevant SARS-CoV-2 variants per ten million cases versus the cumulative number $N$ of cases in the world.

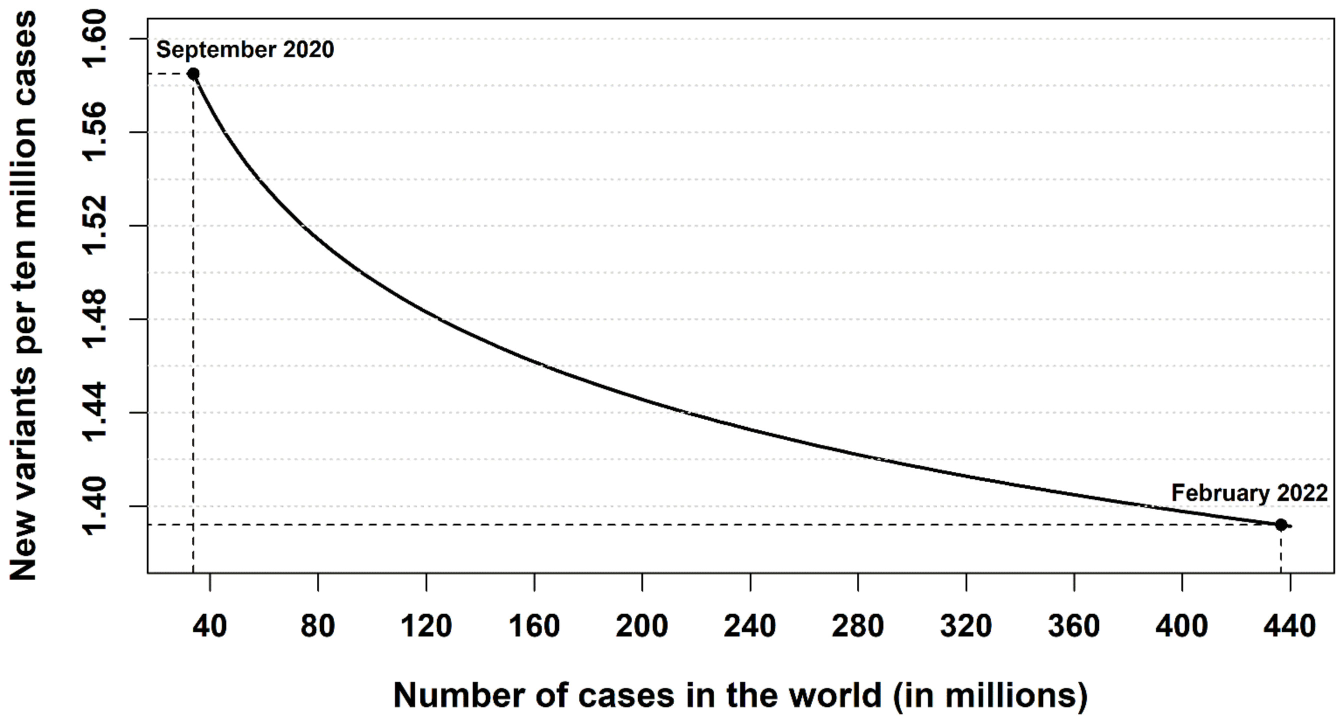


**Figure D2**. Number $n$ of new relevant SARS-CoV-2 variants per ten million cases versus the cumulative number of cases in the world. From September 2020 to February 2022 $n$ decreased from 1.59 to 1.39.

#### References of Appendix D

[D1] ECDC (European Centre for Disease Prevention and Control). SARS-CoV-2 variants of concern. https://www.ecdc.europa.eu/en/covid-19/variants-concern.

[D2] ECDC (European Centre for Disease Prevention and Control). COVID-19 situation update worldwide. https://www.ecdc.europa.eu/en/geographical-distribution-2019-ncov-cases.

[D3] WHO (World Health Organization). Tracking SARS-CoV-2 variants. Working Definitions and Actions Taken. https://www.who.int/en/activities/tracking-SARS-CoV-2-variants/.

### Appendix E

#### Details on the numerical fit

In this Appendix we discuss some details on the fit of WHO data (confidence intervals, residuals and numerical derivatives) and consider the approximation given by the linear regression.

##### CONFIDENCE INTERVALS AND RESIDUALS

The 95% confidence interval (CI) of the constant $k = 1.83 \cdot 10^{-6}$ of the numerical fit of WHO data on relevant SARS-CoV-2 variants is $95\%\ \mathrm{CI} = (1.52 - 2.15) \cdot 10^{-6}$. In the figure below the dashed lines correspond to the upper and lower limits of the 95% CI of the constant $k$.

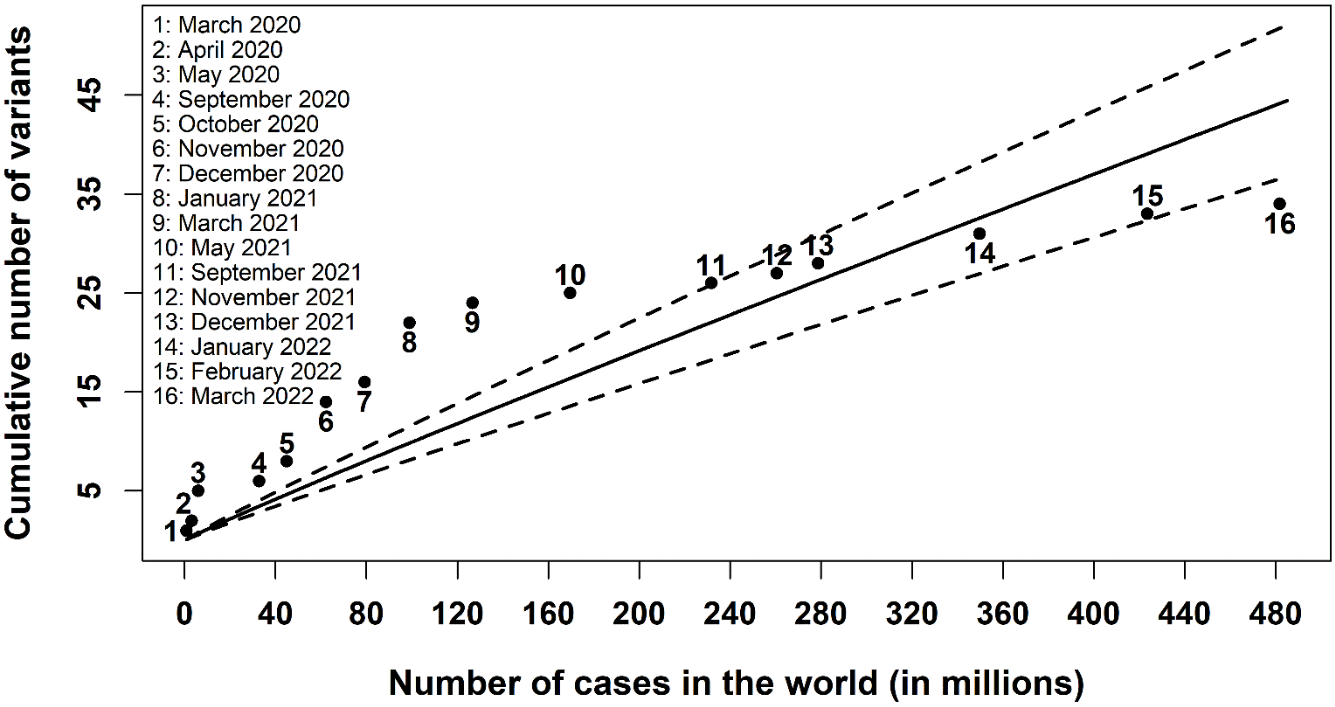


The residual $r_i$ of the $i$-th value in a set of $m$ data is the difference between the observed value $y_i$ and the corresponding value $\hat{y}_i$ predicted by the fit: $r_i = y_i - \hat{y}_i$, with $i = 1, 2, \dots, m$. In our fit the maximum absolute value $r_{max}$ of the residuals $r_i$ is given by

$$r_{max} = \max_i |y_i - \hat{y}_i| = 12.15$$

corresponding to the eighth observation ($i = 8$) shown in the previous figure.

The residual standard deviation $\sigma_r$ is defined as:

$$\sigma_r = \sqrt{\frac{\sum_{i=1}^{m}(y_i - \hat{y}_i)^2}{df}}$$

where $df$ is the degree of freedom: $df = m - p$ ($m$ is the number of observed data and $p$ the number of parameters in the fit: $m = 16$ and $p = 1$ in our case). The residual standard deviation for our fit is

$$\sigma_r = \sqrt{\frac{\sum_{i=1}^{16}(y_i - \hat{y}_i)^2}{16 - 1}} = 6.75$$

The Table below compares the observed and predicted values of relevant variants in each of the sixteen WHO observations between March 2020 and March 2022.

| $i$ | **Earliest detection** | $N$ | **WHO variants** | $v$ (**95% CI**) | $\|r\|$ | $n$ (**95% CI**) |
|---|---|---|---|---|---|---|
| 1 | Mar-2020 | 750890 | 1 | 0.10 (0.08-0.12) | 0.90 | 1.25 (1.04-1.47) |
| 2 | Apr-2020 | 3090445 | 2 | 0.38 (0.31-0.44) | 1.62 | 1.14 (0.95-1.34) |
| 3 | May-2020 | 5934936 | 5 | 0.70 (0.58-0.82) | 4.30 | 1.10 (0.91-1.29) |
| 4 | Sep-2020 | 32730945 | 6 | 3.47 (2.87-4.06) | 2.53 | 1.00 (0.83-1.17) |
| 5 | Oct-2020 | 44888869 | 8 | 4.67 (3.86-5.47) | 3.33 | 0.98 (0.81-1.15) |
| 6 | Nov-2020 | 62195274 | 14 | 6.35 (5.26-7.45) | 7.65 | 0.96 (0.80-1.13) |
| 7 | Dec-2020 | 79231893 | 16 | 7.98 (6.61-9.36) | 8.02 | 0.95 (0.79-1.12) |
| 8 | Jan-2021 | 98925221 | 22 | 9.85 (8.15-11.54) | 12.15 | 0.94 (0.78-1.10) |
| 9 | Mar-2021 | 126697603 | 24 | 12.44 (10.30-14.59) | 11.56 | 0.93 (0.77-1.09) |
| 10 | May-2021 | 169597415 | 25 | 16.40 (13.57-19.23) | 8.60 | 0.92 (0.76-1.07) |
| 11 | Sep-2021 | 231703120 | 26 | 22.04 (18.25-25.84) | 3.96 | 0.90 (0.75-1.06) |
| 12 | Nov-2021 | 260493573 | 27 | 24.63 (20.39-28.88) | 2.37 | 0.90 (0.74-1.05) |
| 13 | Dec-2021 | 278714484 | 28 | 26.26 (21.74-30.79) | 1.74 | 0.89 (0.74-1.05) |
| 14 | Jan-2022 | 349641119 | 31 | 32.57 (26.96-38.18) | 1.57 | 0.88 (0.73-1.04) |
| 15 | Feb-2022 | 423437674 | 33 | 39.06 (32.33-45.79) | 6.06 | 0.88 (0.73-1.03) |
| 16 | Mar-2022 | 481756671 | 34 | 44.15 (36.55-51.76) | 10.15 | 0.87 (0.72-1.02) |

The index $i$ indicates the order of each observation, $N$ is the cumulative number of infected cases in the world, $|r|$ is the absolute value of each residual (difference between observed and predicted values), $v$ (95%CI) is the number of cumulative relevant variants predicted by the model with the corresponding 95% CI and $n$ (95% CI) is the predicted number of new relevant variants per ten million infected cases. The column "WHO variants" lists the number of relevant variants recorded by WHO up to the reported date of detection.

#### NUMERICAL DERIVATIVES

In order to obtain the derivatives of the function underlying the WHO data on relevant SARS-CoV-2 variants, we interpolated the observed data with Wolfram Mathematica by exploiting the B-splines and the Lagrange and Hermite methods. Moreover, we computed the so-called three-points and five-points formulas discussed e.g. by Burden RL and Faires JD in "Numerical analysis" (7th edition. Pacific Grove: Brooks/Cole; 2001).

If $x_0, x_1, \dots, x_q$ are $q+1$ distinct numbers in an interval $I$ and $f(x)$ is a function whose values are known in these points, the $(q+1)$-point formula expressing the derivative $f'$ of the function $f$ in a point $x_j$, with $j = 0, 1, \dots, q$, is given by:

$$f'(x_j) = \sum_{k=0}^{q} f(x_k) \cdot L'_{q;k}(x_j) + \frac{f^{(q+1)}\left(\xi(x_j)\right)}{(q+1)!} \prod_{\substack{k=0 \\ k\neq j}}^{q} (x_j - x_k) \cong \sum_{k=0}^{q} f(x_k) \cdot L'_{q;k}(x_j)$$

where $\xi$ is a point in $I$, depending on $x_j$, and $L'_{q;k}(x)$ is the derivate of the $k$-th coefficient $L_{q;k}(x)$ of the $q$-th Lagrange interpolating polynomial:

$$L_{q;k}(x) = \prod_{\substack{i=0 \\ i\neq k}}^{q} \frac{x - x_i}{x_k - x_i}$$

The most common formulas are those involving three and five evaluating points:

$$f'(x_j) \cong \sum_{\substack{k=j-h \\ k\neq j}}^{j+h} f(x_k) \cdot P'_{h;k}(x_j)$$

where $h = 1$ or $h = 2$ for the three-points or five-points formula, respectively, and

$$P'_{h;k}(x_j) = D\left[\prod_{\substack{i=j-h \\ i\neq j}}^{j+h} \frac{x - x_i}{x_k - x_i}\right]_{x=x_j}$$

being $D[\ \ ]_{x=x_j}$ the derivative with respect to $x$ computed in $x = x_j$.

The sets of three points $\{x_{j-1}, x_j, x_{j+1}\}$ and five points $\{x_{j-2}, x_{j-1}, x_j, x_{j+1}, x_{j+2}\}$, belonging to the complete set $\{x_0, x_1, \dots, x_q\}$, are chosen so that the point $x_j$ where the derivative must be computed is in central position (or as most central as possible), since in this case the approximation error is minimum.

In our fit the set $\{x_0, x_1, \dots, x_q\}$ is given by the $q + 1 = 15 + 1$ points $\{N_1, N_2, \dots, N_{16}\}$ corresponding to the cumulative numbers of infected cases in the world for all the cumulative numbers of relevant variants detected by WHO in the months between March 2020 and March 2022 (see Table 1 of the Results section) and interpolated by the chosen function (B-splines and Lagrange or Hermite polynomials).

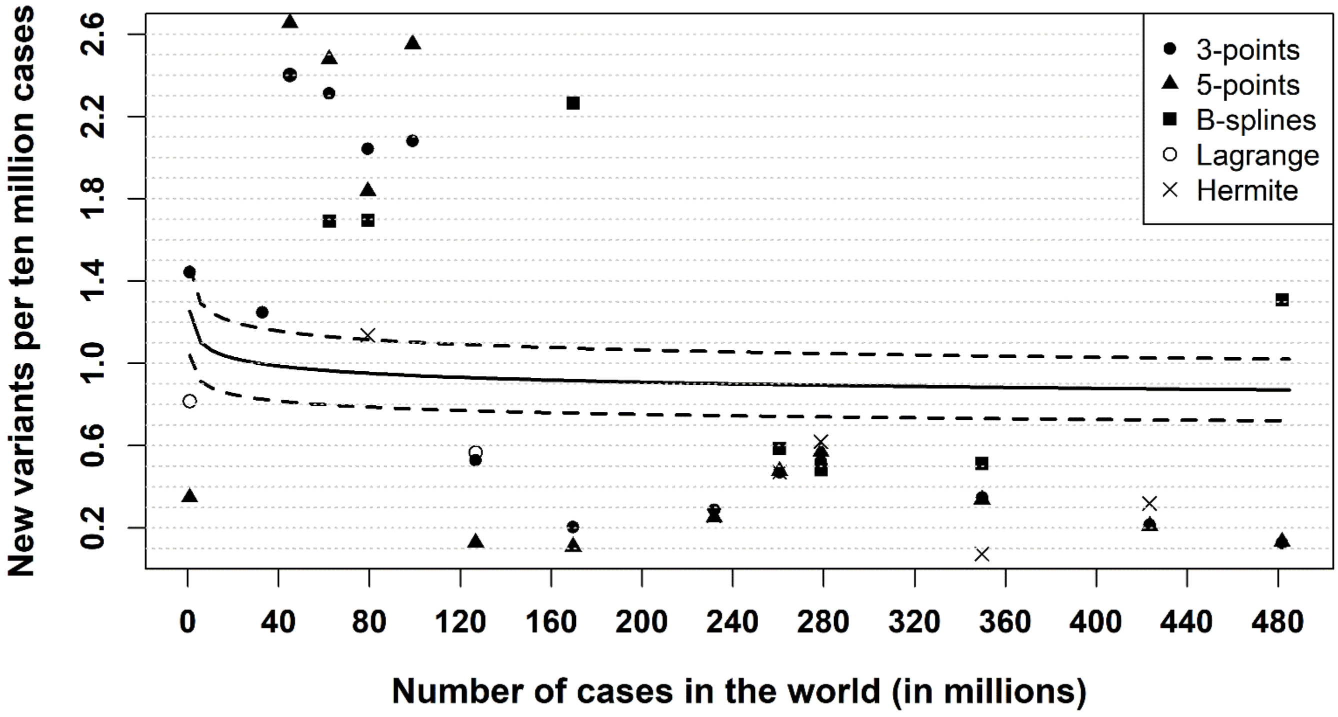


In the previous figure the solid line represents the new variants per ten million cases $n = k \cdot 10^7 \frac{\log N - 1}{(\log N)^2}$, while the dashed lines correspond to the upper and lower limits of the 95% CI $= (1.52 - 2.15) \cdot 10^{-6}$ of the constant $k = 1.83 \cdot 10^{-6}$. The dots represent the new variants per ten million cases computed through the formula $n \cong f'(N) \cdot \Delta N$, where $\Delta N = 10^7$ and the numerical derivative $f'$ of the function $f$ underlying the observed data was obtained with the methods reported in the legend (three or five points formulas, B-splines, Lagrange and Hermite interpolation). Only $n$ values between 0 and 2.6 were considered.

**Linear regression**

The numerical fit $v = k \cdot N / \log N$, where $v$ is the cumulative number of relevant SARS-CoV-2 variants and $N$ is the cumulative number of infected cases, can be approximated by the linear regression $\tilde{v} = h \cdot N$, with $h = 9.27 \cdot 10^{-8}$ and 95% CI = $(7.56 - 10.99) \cdot 10^{-8}$.

The linear model does not satisfy the third condition listed in the Methods section. However, it is close to the logarithmic fit up to large numbers of cases $N$ in the world. Specifically, the difference between the number of relevant SARS-CoV-2 variants predicted by the two fits is zero for $N = 383$ million cases (February 2022), as shown in the figure below, and raises to 10 relevant variants for $N = 1.6$ billion cases in the world (corresponding to about three times the cases recorded from the beginning of the epidemic up to June 2022).

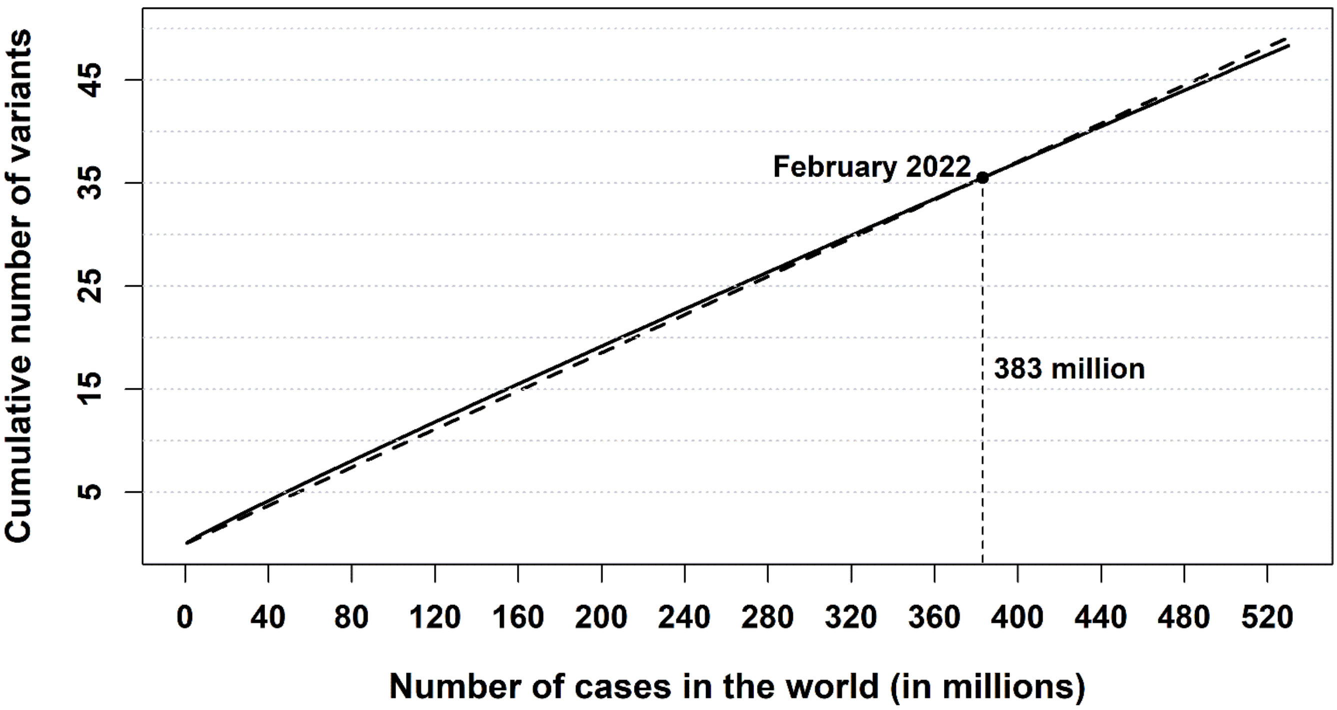


In the logarithmic fit $v = k \cdot N / \log N$ the number $n$ of new relevant variants per ten million cases decreases as the number $N$ of cases increases, as discussed in the Results section: $n = 18.3 \cdot \frac{\log N - 1}{(\log N)^2}$.

On the contrary, in the linear fit $\tilde{v} = h \cdot N$ the number $n$ is constant: $n \cong \tilde{v}'(N) \cdot \Delta N = 0.93$, being $\tilde{v}'(N) = h = 9.27 \cdot 10^{-8}$ and $\Delta N = 10^7$. This different behaviour between the logarithmic and linear fits is represented in the following figure.

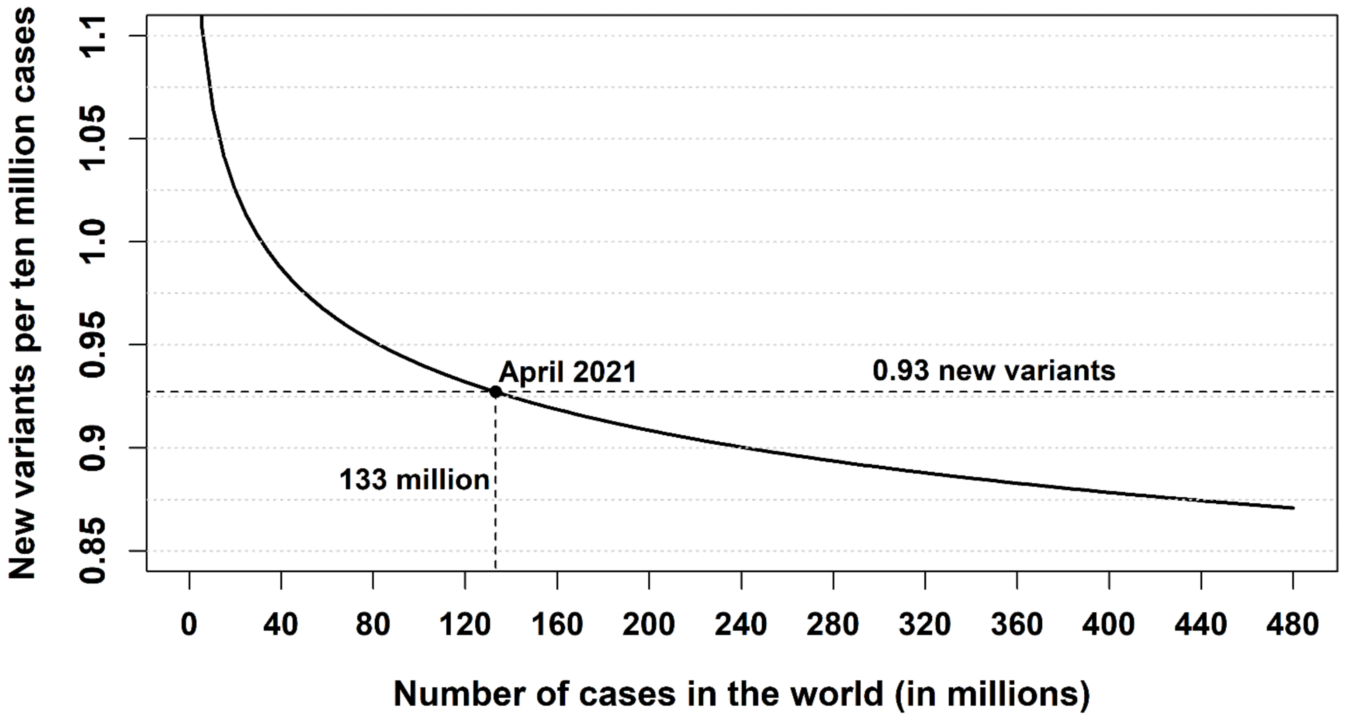


The full circle in the previous figure indicates the intersection of the linear and logarithmic fits, i.e. their common value as well as the corresponding number of infected cases and the date of detection. In the logarithmic fit the number $n$ of new relevant variants per ten million infections decreases – although slowly – as the virus circulates, while in the linear fit $n$ is constant.
